# EEG response during sedation interruption complements behavioral assessment following severe brain injury

**DOI:** 10.1101/2024.10.02.24314815

**Authors:** Charlotte Maschke, Loretta Norton, Catherine Duclos, Miriam Han, Kira Dolhan, Geoffrey Laforge, Allison Frantz, Xiaoyu Wang, Hassan Al-Hayawi, Tianyu Zhang, Raphaël Lavoie, Adrian M. Owen, Stefanie Blain-Moraes

## Abstract

**Background and Objectives:** Accurate assessment of level of consciousness and potential to recover in severe brain injury patients underpins crucial decisions in the intensive care unit but remains a major challenge for the clinical team. The neurological wake-up test (NWT) is a widely used assessment tool, but many patients’ behavioral response during a short interruption of sedation is ambiguous or absent, with little prognostic value. This study assesses the brain’s electroencephalogram response during an interruption of propofol sedation to complement behavioral assessment during the NWT to predict survival, recovery of consciousness, and long-term functional outcome in acute severe brain injury patients.

**Methods:** We recorded 128-channel EEG of 41 severely brain-injured patients during a clinically indicated NWT. The Glasgow Coma Scale (GCS) was used to assess behavioral responsiveness before and after interruption of sedation (GCS_observed_). During the NWT, nine patients regained responsiveness, 13 patients showed ambiguous responsiveness and 19 patients were not responsive. Brain response to sedation interruption was quantified using EEG power, spatial ratios and the spectral exponent. We trained a linear regression model to identify brain patterns related to regaining behavioral responsiveness. We then applied this model to patients whose behavioral responses were ambiguous or absent, using their NWT brain responses to predict a change in behavioral response (ΔGCS_predicted_). Prognostic value of the ΔGCS_predicted_ was assessed using the Mann-Whitney-U test and group-separability. The patients’ survival, recovery of responsiveness, and functional outcomes were assessed up to 12 months post-recording.

**Results:** EEG patterns during interruption of sedation reliably predicted the GCS_observed_ in patients who regained responsiveness during the NWT. Electrophysiological patterns of waking-up were observed in some patients whose behavioral response was ambiguous or absent. Compared to the GCS_observed_, the ΔGCS_predicted_ improved separability of prognostic groups and significantly distinguished patients according to survival (U = 87, p<0.05). The EEG-trained model outperformed outcome predictions of the patients’ attending physician and predictions based on the patients’ APACHE score.

**Discussion:** EEG can complement behavioral assessment during the NWT to improve prognostication, inform clinicians, family members and caregivers, and to set realistic goals for treatment and therapy.

## 1 Introduction

The accurate assessment of severely brain injured patients’ levels of consciousness and potential to recover underpins the most crucial clinical decisions in the intensive care unit (ICU) but remains a major challenge for the clinical team. Behavioral assessments, such as the Glasgow Coma Scale (GCS) ^1^, are widely used to evaluate patients based on their level of arousal, reaction to pain and capacity to follow commands. However, in patients who have suffered moderate to severe brain injury, consciousness and responsiveness can be completely dissociated; in other words, patients can be behaviorally unresponsive despite being covertly conscious ^2–4^.

Sedation plays a key role in the early treatment of acutely brain-injured patients ^5,6^. During the first days of ICU admission, these patients are often continuously sedated to regulate pain and agitation, improve tolerance to intubation and reduce the risk of secondary injury ^5,6^. However, extended periods of sedation also limit the clinical team’s ability to identify changes in the patient’s mental status and to separate injury-related impairments from drug-induced effects ^7^. It has therefore become routine practice to interrupt sedation at least once a day to allow patients to wake up for a neuro-cognitive assessment; this practice is known as the neurological wake-up test (NWT) ^5–7^. During this short sedation interruption, the *presence* of signs of awareness of self and of environment are key indicators for early recovery of consciousness and predictive of a good long-term recovery ^8,9^. However, the *absence* of a behavioral response has less diagnostic or prognostic value, as it may be caused by confounding factors ranging from sensory or motor impairments to pain and fatigue.

Accordingly, a multitude of methods have been developed to assess patients by using electrophysiology and neuroimaging techniques to bypass their behavioral capacity ^10–13^. However, most methods require patients to tolerate an extended period without sedation and are therefore not suited for acute severely brain-injured patients, for whom even a short interruption of sedation may cause increased levels of stress and clinical risk ^14,15^. Our research group has previously shown that the brain’s EEG response to propofol anesthesia *induction* can successfully be used for the assessment of unresponsive patients ^16–19^. We demonstrated that EEG reconfiguration in response to propofol anesthesia has diagnostic ^18^ and prognostic ^17^ value to predict recovery of consciousness in unresponsive patients. However, this protocol ^17^ can only be applied to patients who are not already sedated and thus excludes most severe brain-injury patients during the first days of ICU admission, when many critical treatment decision are made.

In this study, we propose the brain’s EEG response to *interruption* of propofol sedation as a complementary measure to behavioral assessment during the NWT. In particular, EEG power, spatial ratios and spectral exponents have been shown to be reliable markers of propofol-induced unconsciousness ^20–22^. We first hypothesize that we will observe a dynamic change in these EEG features in the patients who regained responsiveness during an interruption of propofol sedation. We further hypothesize that changes in these EEG features can be decoupled from the behavioral response in some patients, with dynamic changes in EEG features despite a lack of behavioral response. Finally, we hypothesize that the EEG response to interruption of sedation provides prognostic value for survival and functional outcome of patients who show *absent* or *ambiguous* behavioral response during the NWT.

Continuous EEG monitoring and, in resource-limited settings, serial EEGs, are routinely used to assess the potential for neurological recovery ^23^. Our approach proposes a novel way of capturing dynamic and reactive electrophysiological features, which is highly translatable to clinical practice and resource- and time-efficient. In current clinical practice, early prognostication of brain-injured patients remains an ‘uncertain art and science’ ^24^; our study aims to support clinical decision making by providing a tool that complements behavioral assessment with neurological markers of return of consciousness.

## 2 Methods

### 2.1 Participants

This study was part of a larger protocol, previously published by Duclos et al. ^25^. Patients were recruited from four different intensive care units in Canada: 1) Montreal General Hospital; 2) Montreal Neurological Institute; 3) London University Hospital and 4) London Victoria Hospital. Since 2020, a total of 1878 patients were screened; 183 patients were eligible for the study; consent was obtained for 53 patients; and EEG data was acquired from 47 patients.

Patients were included if they were at least 18 years old, within 14-days of ICU admission and continuously sedated following a brain injury (e.g., stroke, anoxic injury, traumatic brain injury, subarachnoid hemorrhage). Patients were only recruited if an interruption of sedation was planned as part of the standard of care and if they were deemed medically suitable for the study by their attending intensivist. Patients were excluded according to the following criteria: low comprehension of English or French; injuries which hindered high-density EEG (e.g., large bandages, infections, spine fracture); presence of status epilepticus; and history of pre-existing dementia or mild cognitive impairment ^25^. The study was approved by the Research Ethics Board of the McGill University Health Centre (Project ID 2020-5972) and the Western University Health Science Research Ethics Board (Project ID 114303). Written informed consent was provided by the patient’s legal representative family member, in accordance with the Declaration of Helsinki.

### 2.2 Electroencephalography data acquisition and anesthetic protocol

EEG data was recorded in 5 phases: 1) pre-interruption of continuous sedation; 2) during the transition after interruption of sedation; 3) during a resting state off sedation; 4) during the transition after re-initiation of sedation and 5) resumption of continuous sedation (see Figure 1 for schematic illustration of the recording protocol). Phase 1 was recorded for 10 minutes, the other phases had variable durations (see below). EEG data were recorded using a high-density 128 channel saline net and an Amps 400 amplifier (Electrical Geodesic, Inc., USA). Data were recorded at a sampling rate of 1kHz and were referenced to the vertex; electrode impedances were reduced to below 5KΩ prior to recording.

**Figure 1:**
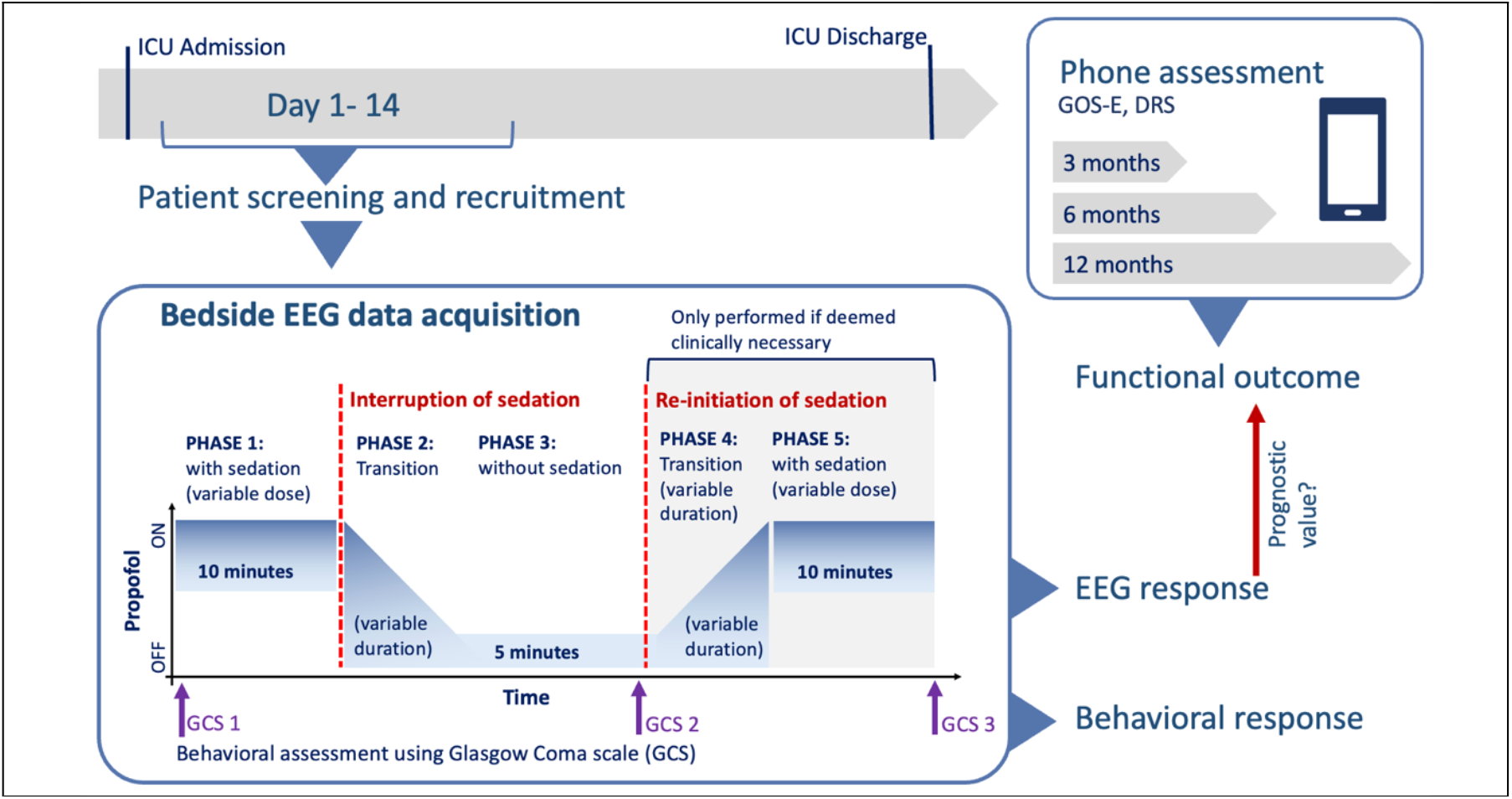
Schematic illustration of study design. Patients were recruited within 14 days of admission to the ICU, and EEG was recorded in in 5 phases: 1) on sedation, 2) during the transition post-interruption of sedation, 3) off sedation, 4) during the transition post re-initiation of sedation and 5) when sedation was on again. Behavioral assessment using the GCS was performed three times during the protocol. Patients’ functional outcome was determined using phone assessments. In this study, we evaluated the value of behavioral and EEG response to prognosticate patients’ long-term functional outcome. ICU: intensive care unit, GCS: Glasgow Coma Scale, GOS-E: Glasgow Outcome Scale -Extended, EEG: electroencephalography

Patient responsiveness was assessed by the attending nurse using the GCS^1^ three times during the protocol: before Phase 1 (GCS_sedation_); at the end of Phase 3 (GCS_observed_); and at the end of Phase 5 (see Figure 1 for schematic illustration of recording protocol).

Parameters for interruption of sedation were not standardized. Instead, EEG response was recorded during routine ICU patient assessments. Thus, the initial sedation concentration, duration of sedation interruption and the need for a re-initialization of sedation were determined for every patient individually by the attending nurse or intensivist according to standard of care. Initial concentrations of propofol infusion varied between 16 to 83 μg/kg/min (see Table 1). The total time between interruption and re-initiation of sedation was determined by the attending nurse based on medical judgement (i.e., patient’s level of agitation, inter-cranial pressure, respiration) and varied between 11 and 33 minutes. Of the 47 patients whose EEG was recorded, 6 patients were excluded due to increased levels of agitation and head movement, which caused excessive noise in the EEG data.

**Table 1:**
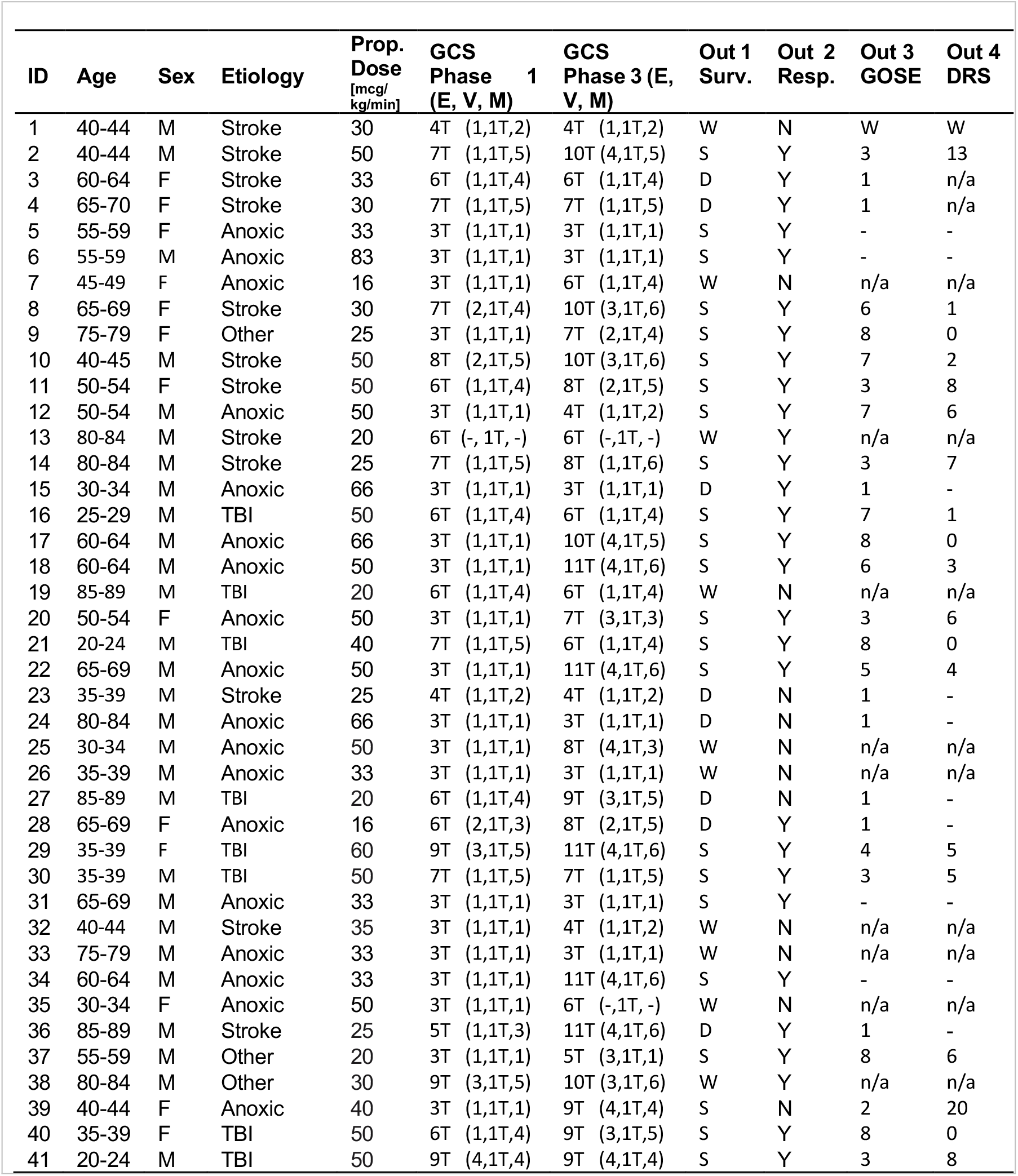
Demographic information of 41 patients included in this study. ID: patient identifier; Sex (M: Male, F: Female), Prop: Propofol concentration in mcg/kg/min, score of Glasgow coma scale (GCS) in Phase 1 and Phase 3 (E: Eye opening, V: verbal Response, M: motor response, T: Intubated); Outcome 1: Survival (W: Withdrawn from life sustaining treatment, S: Survived, D: deceased), Outcome 2: Responsiveness (N: no-did not regain responsiveness, Y: yes-did regain responsiveness), Outcome 3: maximal score on Glasgow outcome scale-extended (GOS-E), Outcome 4: minimal score on Disability Rating Scale (DRS).

| ID | Age | Sex | Etiology | Prop. Dose [mcg/kg/min] | GCS Phase 1 (E, V, M) | GCS Phase 3 (E, V, M) | Out 1 Surv. | Out 2 Resp. | Out 3 GOSE | Out 4 DRS |
| --- | --- | --- | --- | --- | --- | --- | --- | --- | --- | --- |
| 1 | 40-44 | M | Stroke | 30 | 4T (1,1T,2) | 4T (1,1T,2) | W | N | W | W |
| 2 | 40-44 | M | Stroke | 50 | 7T (1,1T,5) | 10T (4,1T,5) | S | Y | 3 | 13 |
| 3 | 60-64 | F | Stroke | 33 | 6T (1,1T,4) | 6T (1,1T,4) | D | Y | 1 | n/a |
| 4 | 65-70 | F | Stroke | 30 | 7T (1,1T,5) | 7T (1,1T,5) | D | Y | 1 | n/a |
| 5 | 55-59 | F | Anoxic | 33 | 3T (1,1T,1) | 3T (1,1T,1) | S | Y | - | - |
| 6 | 55-59 | M | Anoxic | 83 | 3T (1,1T,1) | 3T (1,1T,1) | S | Y | - | - |
| 7 | 45-49 | F | Anoxic | 16 | 3T (1,1T,1) | 6T (1,1T,4) | W | N | n/a | n/a |
| 8 | 65-69 | F | Stroke | 30 | 7T (2,1T,4) | 10T (3,1T,6) | S | Y | 6 | 1 |
| 9 | 75-79 | F | Other | 25 | 3T (1,1T,1) | 7T (2,1T,4) | S | Y | 8 | 0 |
| 10 | 40-45 | M | Stroke | 50 | 8T (2,1T,5) | 10T (3,1T,6) | S | Y | 7 | 2 |
| 11 | 50-54 | F | Stroke | 50 | 6T (1,1T,4) | 8T (2,1T,5) | S | Y | 3 | 8 |
| 12 | 50-54 | M | Anoxic | 50 | 3T (1,1T,1) | 4T (1,1T,2) | S | Y | 7 | 6 |
| 13 | 80-84 | M | Stroke | 20 | 6T (-, 1T, -) | 6T (-, 1T, -) | W | Y | n/a | n/a |
| 14 | 80-84 | M | Stroke | 25 | 7T (1,1T,5) | 8T (1,1T,6) | S | Y | 3 | 7 |
| 15 | 30-34 | M | Anoxic | 66 | 3T (1,1T,1) | 3T (1,1T,1) | D | Y | 1 | - |
| 16 | 25-29 | M | TBI | 50 | 6T (1,1T,4) | 6T (1,1T,4) | S | Y | 7 | 1 |
| 17 | 60-64 | M | Anoxic | 66 | 3T (1,1T,1) | 10T (4,1T,5) | S | Y | 8 | 0 |
| 18 | 60-64 | M | Anoxic | 50 | 3T (1,1T,1) | 11T (4,1T,6) | S | Y | 6 | 3 |
| 19 | 85-89 | M | TBI | 20 | 6T (1,1T,4) | 6T (1,1T,4) | W | N | n/a | n/a |
| 20 | 50-54 | F | Anoxic | 50 | 3T (1,1T,1) | 7T (3,1T,3) | S | Y | 3 | 6 |
| 21 | 20-24 | M | TBI | 40 | 7T (1,1T,5) | 6T (1,1T,4) | S | Y | 8 | 0 |
| 22 | 65-69 | M | Anoxic | 50 | 3T (1,1T,1) | 11T (4,1T,6) | S | Y | 5 | 4 |
| 23 | 35-39 | M | Stroke | 25 | 4T (1,1T,2) | 4T (1,1T,2) | D | N | 1 | - |
| 24 | 80-84 | M | Anoxic | 66 | 3T (1,1T,1) | 3T (1,1T,1) | D | N | 1 | - |
| 25 | 30-34 | M | Anoxic | 50 | 3T (1,1T,1) | 8T (4,1T,3) | W | N | n/a | n/a |
| 26 | 35-39 | M | Anoxic | 33 | 3T (1,1T,1) | 3T (1,1T,1) | W | N | n/a | n/a |
| 27 | 85-89 | M | TBI | 20 | 6T (1,1T,4) | 9T (3,1T,5) | D | N | 1 | - |
| 28 | 65-69 | F | Anoxic | 16 | 6T (2,1T,3) | 8T (2,1T,5) | D | Y | 1 | - |
| 29 | 35-39 | F | TBI | 60 | 9T (3,1T,5) | 11T (4,1T,6) | S | Y | 4 | 5 |
| 30 | 35-39 | M | TBI | 50 | 7T (1,1T,5) | 7T (1,1T,5) | S | Y | 3 | 5 |
| 31 | 65-69 | M | Anoxic | 33 | 3T (1,1T,1) | 3T (1,1T,1) | S | Y | - | - |
| 32 | 40-44 | M | Stroke | 35 | 3T (1,1T,1) | 4T (1,1T,2) | W | N | n/a | n/a |
| 33 | 75-79 | M | Anoxic | 33 | 3T (1,1T,1) | 3T (1,1T,1) | W | N | n/a | n/a |
| 34 | 60-64 | M | Anoxic | 33 | 3T (1,1T,1) | 11T (4,1T,6) | S | Y | - | - |
| 35 | 30-34 | F | Anoxic | 50 | 3T (1,1T,1) | 6T (-, 1T, -) | W | N | n/a | n/a |
| 36 | 85-89 | M | Stroke | 25 | 5T (1,1T,3) | 11T (4,1T,6) | D | Y | 1 | - |
| 37 | 55-59 | M | Other | 20 | 3T (1,1T,1) | 5T (3,1T,1) | S | Y | 8 | 6 |
| 38 | 80-84 | M | Other | 30 | 9T (3,1T,5) | 10T (3,1T,6) | W | Y | n/a | n/a |
| 39 | 40-44 | F | Anoxic | 40 | 3T (1,1T,1) | 9T (4,1T,4) | S | N | 2 | 20 |
| 40 | 35-39 | F | TBI | 50 | 6T (1,1T,4) | 9T (3,1T,5) | S | Y | 8 | 0 |
| 41 | 20-24 | M | TBI | 50 | 9T (4,1T,4) | 9T (4,1T,4) | S | Y | 3 | 8 |

Of the remaining 41 patients, 20 had no medical need for re-initiation of sedation after interruption (i.e., they showed good tolerance to the interruption of sedation or began to obey commands). For these patients, the EEG acquisition was stopped after phase 3 (see Figure 1, see Table 1). For 10 patients, phase 2 (i.e. anesthesia infusion) was not recorded. A total of 21 patients followed the full protocol of 5 states. To include the maximum number of participants, we report the EEG effect of interruption of sedation (i.e., Phase 1-3, n = 41). The same analysis on the EEG effect of re-initialization of propofol (i.e., Phase 3-5, n = 21) is provided in the Supplementary Material.

### 2.3 Data preprocessing

Preprocessing was performed using MNE python ^26^. EEG data was re-referenced to 250 Hz, bandpass-filtered between 0.1 and 50 Hz and Notch-filtered at 60 Hz. Channels with excessive noise and non-physiological artifacts were selected manually and rejected from the data before average referencing. Non-brain electrodes were removed from the subsequent analysis, yielding a maximum of 105 brain channels. Data were segmented in non-overlapping epochs of 10 seconds and evaluated by a trained experimenter. Epochs with non-physiological or movement artifacts were rejected from the subsequent analysis. A total of 113 recordings with an average length of 8.6 minutes and 95 channels were included in the final analysis (see Supplementary Material, Table 1).

### 2.4 Feature extraction

The brain response to the interruption of sedation was estimated using three families of EEG markers: 1) absolute and relative power; 2) spatial ratios; and 3) the spectral exponent. The sum of absolute power of the broadband spectrum and relative power was calculated in five frequency bands: delta (1-4 Hz), theta (4-8 Hz), alpha (8-13 Hz), low beta (13-20 Hz) and high beta (20-30 Hz). Power spectral density between 1 and 45 Hz was estimated using the Welch algorithm with a 2 second window and 50% overlap. Relative power was defined as the area under the power spectrum in the given frequency range, divided by the total area under the full power spectrum, resulting in the percentage contribution of a specific bandwidth to the total power. We estimated the offset and exponential decay of power over frequency using the aperiodic component (referred to as the spectral exponent). The spectral exponent and offset were estimated using the ‘fitting oscillation and one over f’ algorithm ^27^ in the 1-45 Hz range (min_peak_height=0.1, max_n_peaks=3).

All features were calculated on each 10-second epoch and channel individually. We then summarized features by extracting 1) the space-averaged feature strength, 2) the standard deviation of the feature over space, and 3) the posterior-anterior ratio (i.e., the geometric mean of power in posterior electrodes, divided by geometric mean of power in anterior electrodes), with each method yielding one value per epoch.

### 2.5 EEG Dynamic Change

The change in each EEG feature induced by the interruption of propofol was defined as the difference between each feature’s Phase 1 average and Phase 3 average (See Figure 2, amount of change). To evaluate whether this difference was induced by the interruption of sedation rather than spontaneous fluctuations, we assessed the proportion of datapoints post-interruption (i.e., Phase 2 and 3) that were one standard deviation above or below the Phase 1 features (see Figure 2, certainty of change). The resulting value can be interpreted as a ‘certainty score’, with values close to 1 indicating high certainty that the respective feature changed after interruption of sedation, and values of 0 indicating that changes were driven by natural fluctuations in the EEG over time. For the patients for whom Phase 2 was not recorded, certainty was calculated using data from Phase 3 only.

**Figure 2:**
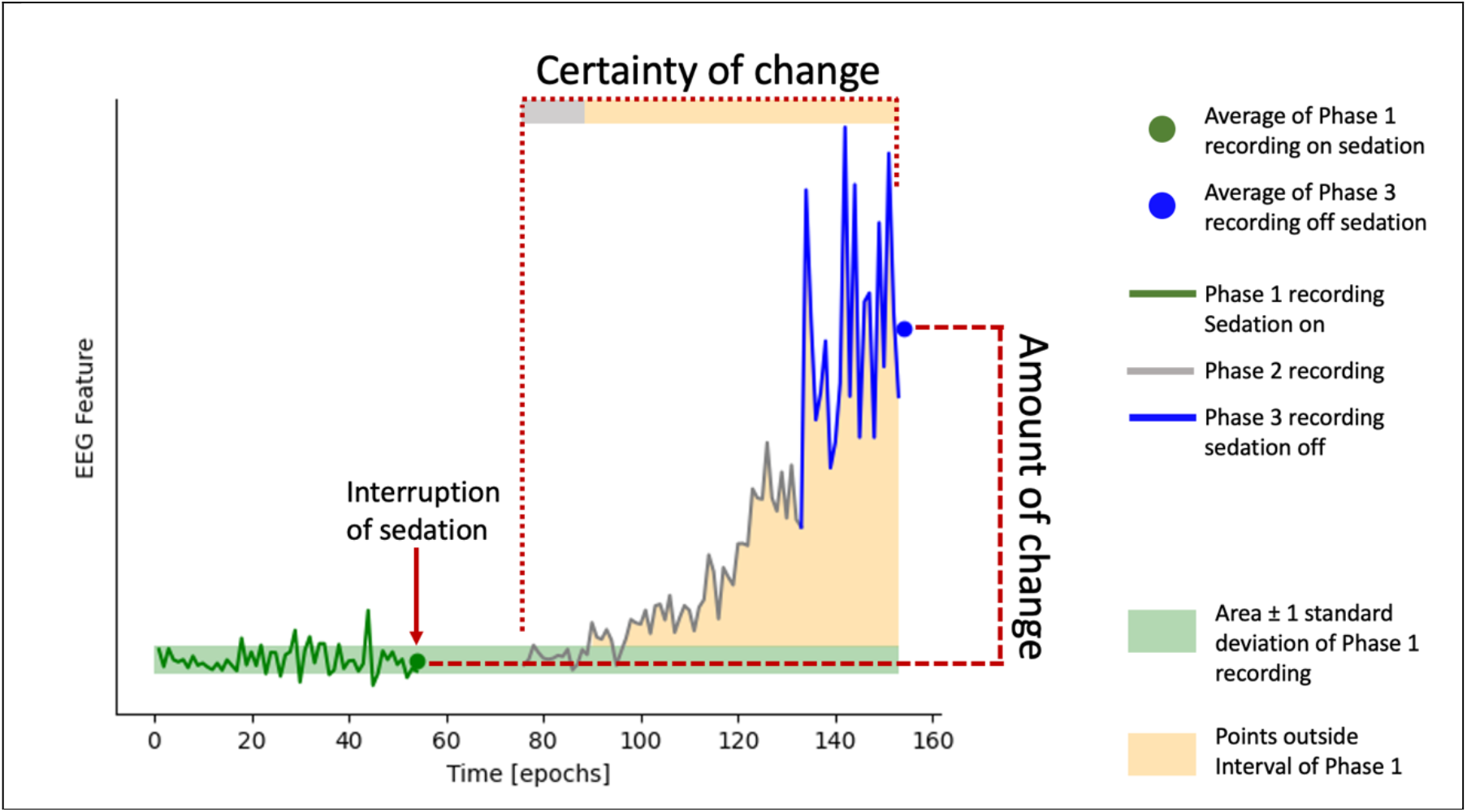
Illustration of EEG feature selection. Colored line represents an example EEG feature (illustrated here is total EEG power over all frequency bands). Green, grey and blue lines indicate values of phase 1, 2 and 3, averaged over electrodes, respectively. The green shaded area indicates the area of ± 1 standard deviation around the mean of Phase 1 signal. The orange shaded area indicates all recording points of Phase 2 and 3 which reached values outside this range. The amount of change was determined by the mean average between Phase 3 and Phase 1. Certainty was rated as the percentage of points outside the green shaded area.

The amount of change in each feature was weighted by the certainty score (i.e., amount of change *certainty of change). This dynamic change score was calculated individually on the mean, standard deviation and posterior-anterior ratio of each feature (i.e., total power, delta, theta, alpha, low beta, high beta, exponent and offset), yielding a total of 24 features (i.e., 8 mean, 8 standard deviation, 8 posterior-anterior ratio). We present all analyses on EEG recordings during interruption of sedation (i.e., Phase 1-3) in the main manuscript; the analysis was reproduced for the re-initiation of sedation (i.e., Phase 3-5) and described in the Supplementary Material (see Supplementary Methods)

### 2.6 Grouping according to behavioral response

Participant behavioral responsiveness was assessed using the GCS before, during and after the interruption of sedation (see Figure 1). The change in behavioral responsiveness (ΔGCS) during the NWT was defined as the difference between GCS score off sedation and pre-interruption of sedation: ΔGCS = GCS_observed_ – GCS_sedation_.

ΔGCS and GCS_observed_ were used to categorize patients into three groups: Nine patients regained ability to obey commands during the NWT and were assigned to ‘Group A: regained responsiveness’ (see Figure 3, green markers, see Table 1). Thirteen patients showed increased levels of behavior during interruption of sedation (i.e., ΔGCS Δ 2), despite not obeying commands and were categorized as ‘Group B: Some behavioral change’ (see Figure 3, orange markers). Nineteen patients did not show any or only minimal behavioral change during NWT (i.e., ΔGCS < 2) and were assigned to ‘Group C: No behavioral change’ (see Figure 3, red markers).

**Figure 3:**
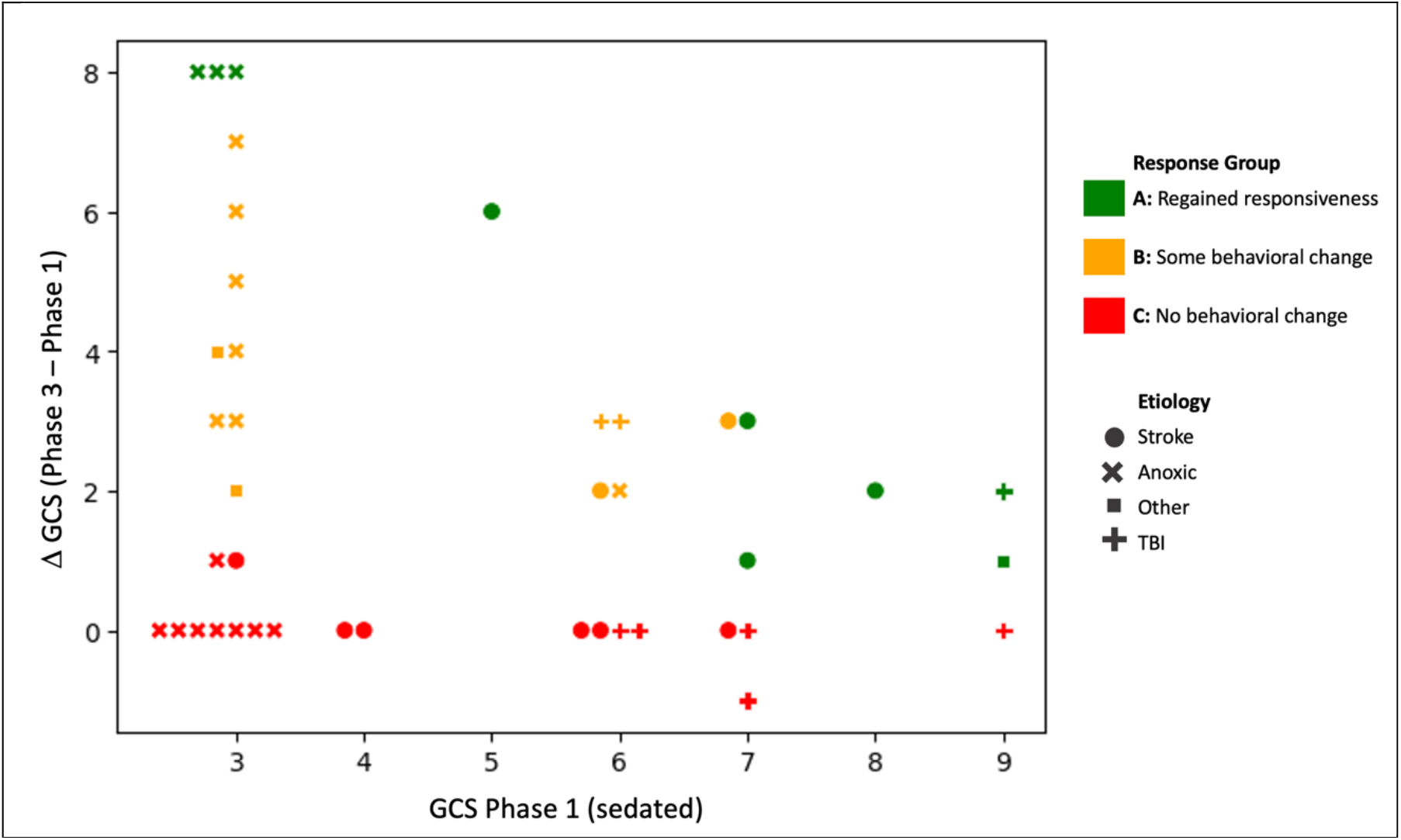
Behavioral response groups. Summary of 41 patients’ GCS score before interruption of sedation and difference in GCS during interruption of sedation (ΔGCS). Each marker indicates a single patient, colored by response group. Marker shape encodes etiology. All participants were intubated at the time of assessment. GCS: Glasgow Coma Scale, TBI: Traumatic Brain Injury

### 2.7 Machine learning analysis

We developed a machine learning model (a Linear Ridge regression trained using scikit learn ^28^) that used the EEG dynamic change score to predict the ΔGCS of Group A participants. For the training, participants’ age, concentration of sedation and time without sedation were combined with the 24 EEG features. Model performance was evaluated using mean absolute error (i.e., average error between true and predicted ΔGCS) and a leave-one participant out cross validation. A good model performance indicates that the model has learned to identify EEG patterns which are related to behavioral responsiveness during the NWT.

We then used the model to predict the change in GCS (ΔGCS_predicted_) during the NWT in participants who showed ambiguous or absent behavioral response (Group B and C). Behavioral responsiveness can be dissociated from the cognitive presence of consciousness (i.e. covert consciousness) ^2–4^. We therefore did not expect to see a high prediction accuracy for Group B and C. Instead, a high deviation between ΔGCS and ΔGCS_predicted_ could indicate that participants show electrophysiological signs of waking-up despite absent or ambiguous behavioral responsiveness.

### 2.8 Outcome assessment and prognostic analysis

Functional outcome was assessed at 3, 6 and 12 months post-EEG recording using a phone assessment of the Glasgow Outcome scale-extended (GOS-E) ^29,30^ and the Disability Rating Scale (DRS) ^31^. Depending on the participant’s functional capabilities, questions were answered by the participant or participant’s next of kin. Four follow-up calls could not be completed due to changed contact information and inability to reach patients’ next of kin. We quantified functional outcome as the maximal GOS-E score (i.e., 0 indicating death) and minimal DRS score (i.e., 0 indicating no disability) reached within one-year post-EEG. In the case of death, we additionally asked whether the participant regained capacity to obey commands at any time prior to death.

As this study aimed to investigate the value of EEG in patients with *absent* or *ambiguous* behavioral responses, the analysis of prognostic value was only performed on participants from Group B and C (n = 32). Group A was used for model training and was not included in the prognostic analysis.

Prognostic analysis was performed based on three outcome criteria: 1) survival; 2) recovery of responsiveness; and 3) functional outcome (i.e., GOS-E and DRS). For the prognostication of survival, 23 patients were included (16 survived, 7 died within three months post-recording). Due to uncertainty of the natural progression of their recovery, participants who had a withdrawal of life sustaining treatment (WLST) were excluded from this analysis (n = 9, see Table 2). For the prognostication of recovery of responsiveness, 24 patients were included (20 regained responsiveness, 4 did not). Participants who had a WLST and had never regained responsiveness (n=8) were excluded from this analysis. For the prognostication of functional outcome, 20 participants were included (9 WLST excluded, 3 missed follow-ups, see Table 2).

**Table 2:** Summary of outcome measures, split by response groups. GOS-E: Glasgow Outcome Scale-Extended, DRS: Disability rating score, N: Number of participants, WLST: withdraw of life-sustaining treatment

|  | N | N<br>Regained<br>responsiveness | N<br>Did not regain<br>responsiveness | N<br>Follow-up<br>calls | GOS-E | DRS |
| --- | --- | --- | --- | --- | --- | --- |
| **Group A (n = 9)** |  |  |  |  |  |  |
| Survivors | 7 | 7 | 0 | 6<br>(1 missed) | Mean: 5.16<br>Std: 1.34<br>Range: 3 - 7 | Mean: 3.67<br>Std: 1.97<br>Range: 1 - 7 |
| Non-survivors | 1 | 1 | 0 | - | GOSE = 1 | N.A. |
| WLST | 1 | 1 | 0 | - | - | - |
| **Group B (n = 13)** |  |  |  |  |  |  |
| Survivors | 8 | 7 | 1 | 8 | Mean: 5.37<br>Std: 2.64<br>Range: 2 - 8 | Mean: 6.63<br>Std: 6.65<br>Range: 0 - 20 |
| Non-survivors | 2 | 1 | 1 | - | GOSE = 1 | N.A. |
| WLST | 3 | 0 | 3 (excluded) | - | - | - |
| **Group C (n = 19)** |  |  |  |  |  |  |
| Survivors | 8 | 8 | 0 | 5<br>(3 missed) | Mean: 5.6<br>Std: 2.15<br>Range: 3 - 8 | Mean: 4.0<br>Std: 3.03<br>Range: 0 - 8 |
| Non-survivors | 5 | 3 | 2 | - | GOSE = 1 | N.A. |
| WLST | 6 | 1 | 5 (excluded) | - | - | - |

### 2.9 Outcome prediction by attending physicians

To compare our proposed tool to bedside clinical practice, we collected clinicians’ prognostication of participant functional recovery. Attending physicians were asked to predict their patient’s 6- and 12-month GOS-E score. They were also asked to rank their confidence in their prediction on a scale from 0 to 4, with 0 indicating not confident and 4 being very confident. Outcome prediction questionnaires were completed between 1 and 24 hours after the EEG recording. Patient’s APACHE score ^33^ was calculated by a trained clinician. For three patients, an APACHE score could not be determined due to missing clinical information.

### 2.10 Statistical analysis

The prognostic value of the EEG model to predict survival and recovery of responsiveness was performed using a Python implementation of the one-sided Mann-Whitney-U test ^34^, with the expectation of a larger ΔGCS_predicted_ for patients with a favorable outcome. In addition, we defined an optimal decision-threshold by identifying the value which maximized the distance between the favorable and unfavorable group. Using this linear split, we defined values of accuracy, specificity and sensitivity of the group-separability. It is important to note that the machine learning model from the first part of this study only predicted an expected ΔGCS, but not patient outcome. The purpose of the optimal threshold was solely to compare how clinically informative the EEG-predicted response was, compared to the observed behavioral response. The prognostic value for functional outcome was assessed using a Spearman-rank test ^34^.

## 3 Results

### 3.1 EEG features can be decoupled from participant behavioral response

Using the Linear Ridge Regression, the ΔGCS of participants in Group A was predicted from the EEG model with a mean absolute error of 1.57. The most important features for the prediction were the relative power in the low beta, theta and alpha bandwidth and the magnitude and posterior-anterior ratio of the spectral exponent.

The ΔGCS_predicted_ for Groups B and C had an overall error of 2.15. Model prediction for Group B varied between -0.69 and 8.35; model prediction for Group C varied between -0.88 and 8.06. High values of ΔGCS_predicted_ indicate the presence of electrophysiological patterns of waking up, similar to participants with high ΔGCS in Group A. Negative values indicate EEG dynamic changes which were even weaker than the training example with the lowest ΔGCS.

Figure 4 illustrates the predicted and observed ΔGCS. Although the predicted difference of most Group B participants matched the observed difference, some participant’s ΔGCS_predicted_ were highly under- or over-estimated. We use four case examples of the maximally under- and over-estimated ΔGCS_predicted_ to elucidate the different types of brain responses during the NWT. Case 1 and 2 (Patient 33 and 11 in Table 1) showed an absent (ΔGCS = 0) or ambiguous (ΔGCS = 2) behavioral response during the NWT. However, based on the EEG reaction to interruption of sedation, the model generated a ΔGCS_predicted_ of 8.06 and 8.53, respectively. In contrast, Case 3 and 4 (Patient 4 and 7 in Table 1) showed an absent (ΔGCS = 0) or ambiguous (ΔGCS = 3) response during the NWT. In both cases, the model generated a negative ΔGCS_predicted_.

**Figure 4:**
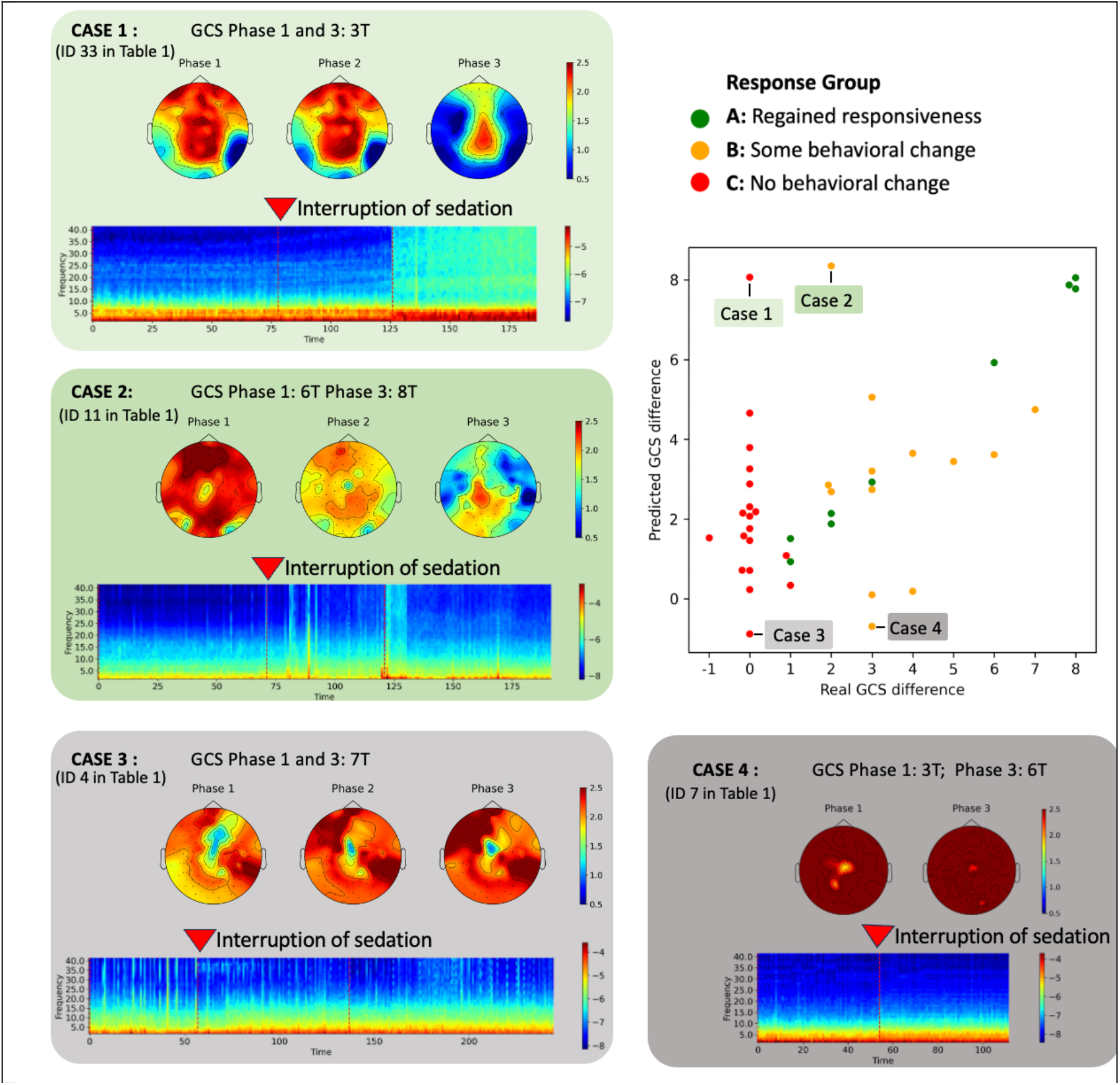
Case examples of contradicting EEG and GCS response. (Top Right) Observed GCS difference (i.e. GCS off sedation – GCS on sedation) and GCS difference predicted by the model. Colors indicate the previously defined response groups. For four highlighted and annotated cases, the EEG response to interruption of sedation is visualized. Each box visualizes a time-resolved spectrogram and topographic maps of spectral exponent for the highlighted case. Dotted lines in the spectrogram indicate the beginning of a new recording phase. In case 4, phase 2 was not recorded. Red arrows indicate the moment when propofol sedation is interrupted. GCS: Glasgow Coma Scale

The time-resolved spectrogram and topographic maps of spectral exponent for the four highlighted cases reveal electrophysiological patterns underpinning the prediction (see Figure 4). While Cases 1 and 2 show a prominent increase of global power and flattening of the spectral exponent following sedation interruption, no such effect was observed for Cases 3 and 4.

Altogether, these cases demonstrate that dynamic EEG changes reveal heterogeneity in the group of participants whose response to the NWT was ambiguous or absent. Despite remaining completely unresponsive, some participants showed EEG patterns that have previously been associated with an increased behavioral response. Our results indicated that EEG features can be decoupled from behavioral responsiveness during the NWT.

### 3.2 EEG response to interruption of sedation has prognostic value for participants with absent or ambiguous behavioral responsiveness

Of the 23 participants included in the prognostic analysis of survival, only 11 had an observed ΔGCS above zero during the NWT. The observed ΔGCS had low predictive value for survival, with an accuracy of 0.61, specificity of 0.56 and sensitivity of 0.71 (Figure 5A). Participants’ maximum observed GCS score was also poorly predictive of survival, with an accuracy of 0.61, specificity of 0.62 and sensitivity of 0.57 (Figure 5B). In contrast, the ΔGCS_predicted_ had higher prognostic value, with an accuracy of 0.74, specificity of 0.75 and sensitivity of 0.71 (Figure 5A). The predicted maximal GCS only minimally increase group separability, with an accuracy of 0.65, specificity of 0.69 and sensitivity of 0.57 (Figure 5B). Observed GCS scores did not significantly differentiate participants according to their outcome; in contrast, the EEG model generated significantly higher ΔGCS_predicted_ for participants who survived (U = 87, p<0.05) (Figure 5A). Supplementary Figure 1 visualizes this effect for each response group individually.

**Figure 5:**
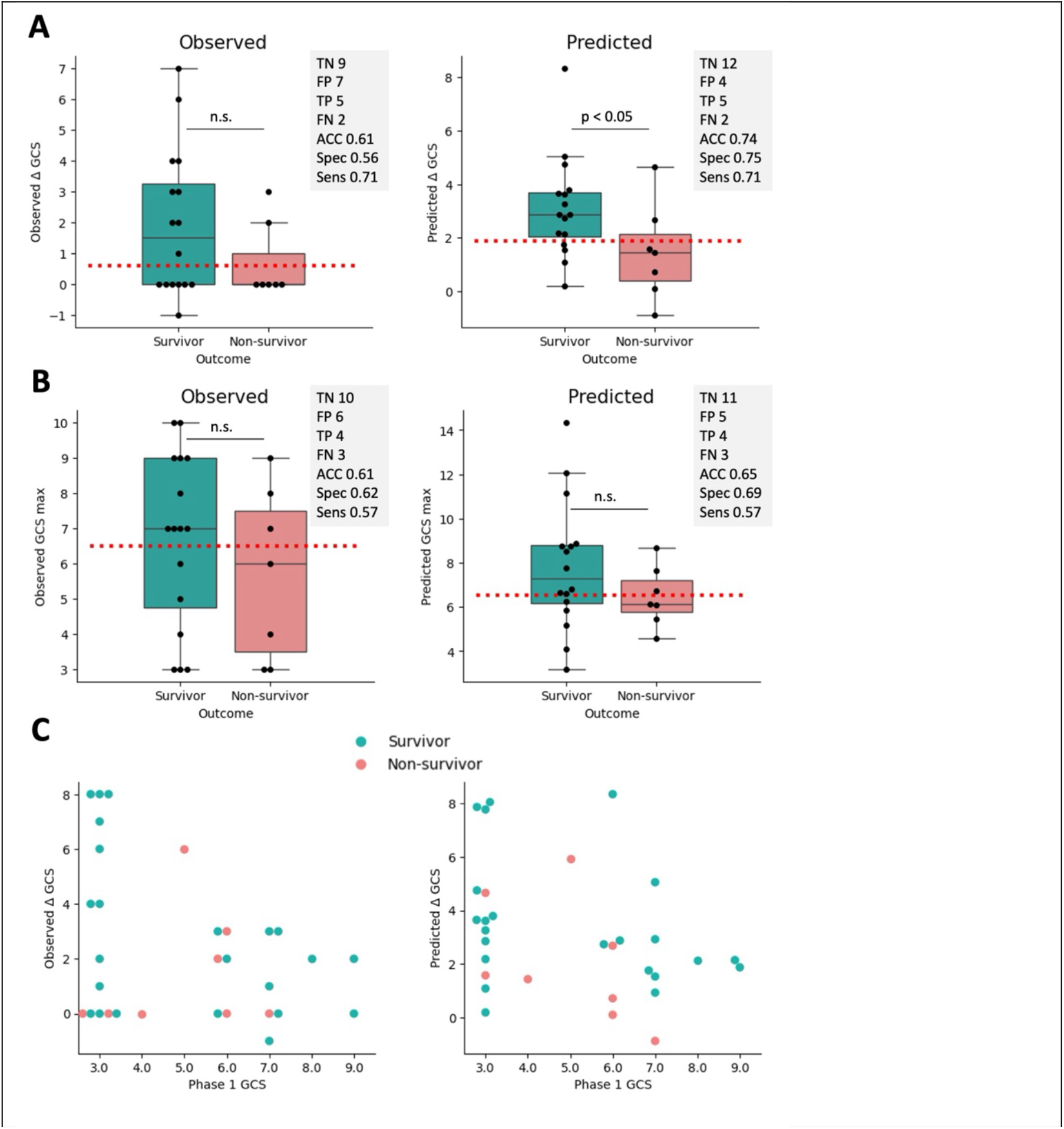
Comparison of observed and EEG-predicted behavioral response. **A)** Observed (left) and model-predicted (right) difference in Glasgow Coma Scale (GCS) during neurological wake-up test, split by patient’s outcome. **B)** Observed (left) and model-predicted (right) maximal Glasgow Coma Scale (GCS) during neurological wake-up test; The dotted line represents the value which best separates the favorable and unfavorable group. The grey box indicates performance matrices of group-separation based on this threshold (TN: true negative, FN: false negative, TP: true positive, FP: false positive, with positive indicating non-survivors, ACC: accuracy, Spec: specificity, Sens: sensitivity). **C)** Visualization of group-separability based on the behaviorally observed response on sedation (i.e. GCS Phase 1) and the observed (left) and model-predicted (right) gain in GCS during NWT.

The prognostic value for recovery of responsiveness remained unchanged between GCS_observed_ and ΔGCS_predicted_ (Supplementary Figure 2). No significant correlation was identified between observed or predicted ΔGCS and maximal GOS-E and DRS score within one -year post-recording. The analysis was repeated using a separate model trained on the brain response to induction of sedation (i.e., Phase 3-5, n = 21) (see Supplementary Methods). Although the effects are reduced due to the smaller sample size, there is a better prognostic separability for survival and recovery of responsiveness using the dynamic EEG changes compared to the observed behavior (Figure S3).

Altogether, we demonstrated that the EEG response following interruption of sedation has prognostic value for survival of participants with absent or ambiguous behavioral responsiveness during the NWT. Based on the EEG response, participants who survived showed a higher ΔGCS_predicted_ compared to participants who did not. All analyses were repeated using only the averaged EEG features from Phase 1 and Phase 3 independently. There was no increase in prognostic value using the EEG from Phase 1 (Figure S3), though the EEG from Phase 3 had prognostic value for recovery of responsiveness (Figure S3). Figure 5C clearly illustrates the improvements offered by a combined neuro-behavioral assessment during the wake-up test.

### 3.3 Prognostic value of EEG response to interruption of sedation could benefit clinical decision making

Physicians’ predicted GOS-E score at 6 months positively correlated with participants’ GCS_observed_ (rho(28)= 0.39, p < 0.05, partial correlation corrected by age). As expected, a patient’s behavioral responsiveness had a strong influence on the physician’s prediction of long-term functional outcome, with more responsive patients correlated with predictions of better recovery.

Physicians rated their predictions with a confidence of 1.82 ± 0.71, with 1 indicating ‘slightly confident’ and 2 being ‘fairly confident’. The lowest average confidence score (1.8 ± 0.35) appeared in Group B, who showed an ambiguous behavioral response. Overall, there was a high level of uncertainty in the functional outcome of unresponsive patients, which was maximal when patients show an ambiguous behavioral response.

The absolute distance between the real and the physician-predicted GOS-E score was 1.80 ± 1.42 (Supplementary Material, Figure S4). However, when binarizing patient outcomes according to survival (GOS-E > 1) or recovery of responsiveness (GOS-E > 2), physician predictions could not separate good from poor outcomes (Supplementary Material, Figure S5). Similarly, the APACHE score achieved a maximal prognostic accuracy of 0.37 for survival and 0.62 for recovery of responsiveness (Supplementary Material, Figure S5). Our results confirm that prognostication after brain injury in the tested sample is accompanied by an overall high uncertainty and suggests a tendency to overestimate functional outcome.

## 4 Discussion

In this study, we demonstrated that the EEG dynamic change during the NWT can improve prognostication of severely brain injured patients who show an absent or ambiguous behavioral response to interruption of sedation. The EEG response to interruption of sedation showed electrophysiological signs of waking-up despite behavioral unresponsiveness in some patients. Moreover, the EEG dynamic change during the NWT predicted patient survival better than behavioral responses alone. This has the potential to improve clinical prognostication, as the attending physician’s outcome prediction was highly dependent on the patient’s observed behavior. Assessing covert neurophysiological function alongside behavior can improve clinician confidence in neuroprognostication and decision-making in acute care settings for severely brain injured patients.

Of the four case examples highlighted in the results, the EEG of two changed dynamically in response to interruption of sedation despite the absence of a behavioral response. The first patient suffered an anoxic brain injury and showed a strong EEG dynamic change during the NTW, while his GCS remained constant at 3T. However, these results are inconclusive, as this patient was withdrawn from life sustaining treatment and the natural course of his recovery remains unknown. The second case patient suffered a stroke and had an ambiguous behavioral response during the NWT. Her EEG had strong neurophysiological signs of waking up, which aligned with her survival and recovery of consciousness. The EEG of the third and fourth case examples remained static during the NWT. The third case was a stroke patient who passed away in palliative care but regained the capacity to follow commands in the ICU; the fourth suffered an anoxic brain injury and was withdrawn from life sustaining treatment. This case series illustrates the potential benefits, but also the limitations of the proposed tool. Although the EEG response during the NWT may capture the brain’s capacity for recovery, patient survival and long-term functional outcome are also determined by factors including age, organ function, treatment availability, social support and the occurrence of secondary medical events unrelated to the original brain injury. The technique proposed in this study is thus not intended to be used in isolation, but rather to complement current state-of-the-art medical assessments to generate a better informed prognosis.

The results of this study need to be considered in light of several limitations. First, while all EEG was recorded during a complete interruption of propofol, other medications including morphine, fentanyl (4 patients), midazolam (3 patients), Keppra (2 patients) and haloperidol (2 patients) were not discontinued during the NWT. Second, before the NWT, patients were continuously sedated for durations ranging from hours to days and were exposed to different concentrations of propofol. We accounted for this potential confound in our analysis by adding the concentration of propofol as a feature to the linear regression. No significant difference was found between the concentration of propofol and participant survival or recovery of responsiveness. Third, EEG markers of consciousness are highly variable across etiologies ^20,35^, and this study analyzed brain-injured participants from anoxic, traumatic and stroke etiologies as a single group. However, our study investigated the propofol-induced *relative* change in EEG features, using a within-subject comparison to account for the specifics in each participant’s brain injury. Fourth, the effect of anesthesia on the brain and behavior differs across sex and age ^36–38^. We accounted for age-related differences by including age as a feature in the machine learning model, however, a larger dataset is necessary to identify relevant EEG features in different populations. Fifth, we only extracted features based on fixed EEG bandwidths using a 128-channel EEG system. A larger feature space that accounts for the raw spectrogram over time could be implemented on a larger sample of patients. Finally, all features extracted from the EEG were calculated on a single-channel level. It remains to be validated whether EEG features of clinal EEG systems would be sufficient to provide similar results.

## Data Availability

Data produced in the present study are available upon reasonable request to the authors

## 5 Acknowledgments

This study was funded by a McGill/Western Collaboration Grant from Healthy Brain for Healthy Lives and BrainsCAN (1a-5a-01) and by the Canadian Institutes of Health Research (CIHR, Project Grant no. 480995).

CM is funded through Fonds de recherche du Québec – Santé (FRQS). This research is supported in part by the FRQNT Strategic Clusters Program (2020-RS4-265502 - Centre UNIQUE - Union Neurosciences & Artificial Intelligence – Quebec) (awarded to CM).

AMO is supported by the Canadian Institutes of Health Research (CIHR, #408004) and is a Fellow of the CIFAR Brain, Mind, and Consciousness program.

CD is funded through an FRQS Junior 1 Scholar Award, and Discovery Grant from the Natural Sciences and Engineering Research Council of Canada (NSERC), a CIFAR Global Scholars award, and institutional start-up funding from the Research Centre of the Centre intégré universitaire de santé et de services sociaux (CIUSSS) du Nord-l’Île-de-Montréal.

SBM is funded by a Canada Research Chair (Tier II) and a NSERC Discovery Grant (RGPIN-2023-03619).

## References

1. Teasdale G, Jennett B. Assessment of coma and impaired consciousness : A practical scale. The Lancet. 1974;304(7872):81–84. doi:10.1016/S0140-6736(74)91639-0

2. Monti MM, Vanhaudenhuyse A, Coleman MR, et al. Willful Modulation of Brain Activity in Disorders of Consciousness. New England Journal of Medicine. 2010;362(7):579–589. doi:10.1056/NEJMoa0905370

3. Owen AM, Coleman MR, Boly M, Davis MH, Laureys S, Pickard JD. Detecting Awareness in the Vegetative State. Science. 2006;313(5792):1402–1402. doi:10.1126/science.1130197

4. Sanders RD, Tononi G, Laureys S, Sleigh JW, Warner DS. Unresponsiveness ≠ Unconsciousness. Anesthesiology. 2012;116(4):946–959. doi:10.1097/ALN.0b013e318249d0a7

5. Oddo M, Crippa IA, Mehta S, et al. Optimizing sedation in patients with acute brain injury. Crit Care. 2016;20(1):128. doi:10.1186/s13054-016-1294-5

6. Oddo M, Steiner L. Sedation and analgesia in the neurocritical care unit. In: Oxford Textbook of Neurocritical Care. Oxford University Press; 2016.

7. Kress JP, Pohlman AS, O’Connor MF, Hall JB. Daily Interruption of Sedative Infusions in Critically Ill Patients Undergoing Mechanical Ventilation. New England Journal of Medicine. 2000;342(20):1471–1477. doi:10.1056/NEJM200005183422002

8. Edlow BL, Fecchio M, Bodien YG, et al. Measuring Consciousness in the Intensive Care Unit. Neurocrit Care. 2023;38(3):584–590. doi:10.1007/s12028-023-01706-4

9. Giacino JT. The vegetative and minimally conscious states: consensus-based criteria for establishing diagnosis and prognosis. NeuroRehabilitation. 2004;19(4):293–298.

10. Casali AG, Gosseries O, Rosanova M, et al. A Theoretically Based Index of Consciousness Independent of Sensory Processing and Behavior. Science Translational Medicine. 2013;5(198):198ra105–198ra105. doi:10.1126/scitranslmed.3006294

11. Kazazian K, Abdalmalak A, Norton L, et al. Functional near-infrared spectroscopy as a tool to assess residual and covert consciousness in the intensive care unit (P11-7.003). Neurology. 2023;100(17_supplement_2):2010. doi:10.1212/WNL.0000000000202261

12. Kazazian K, Kolisnyk M, Norton L, Gofton T, Debicki D, Owen A. Predicting the recovery of consciousness after severe acute brain injury using resting state networks: a machine learning approach (N3.001). Neurology. 2023;100(17_supplement_2):2637. doi:10.1212/WNL.0000000000202688

13. Norton L, Kazazian K, Gofton T, et al. Functional Neuroimaging as an Assessment Tool in Critically Ill Patients. Annals of Neurology. 2023;93(1):131–141. doi:10.1002/ana.26530

14. Marklund N. The Neurological Wake-up Test—A Role in Neurocritical Care Monitoring of Traumatic Brain Injury Patients? Front Neurol. 2017;8. doi:10.3389/fneur.2017.00540

15. Skoglund K, Enblad P, Hillered L, Marklund N. The neurological wake-up test increases stress hormone levels in patients with severe traumatic brain injury*. Critical Care Medicine. 2012;40(1):216. doi:10.1097/CCM.0b013e31822d7dbd

16. Blain-Moraes S, Boshra R, Ma HK, et al. Normal Brain Response to Propofol in Advance of Recovery from Unresponsive Wakefulness Syndrome. Front Hum Neurosci. 2016;10. doi:10.3389/fnhum.2016.00248

17. Duclos C, Maschke C, Mahdid Y, et al. Brain Responses to Propofol in Advance of Recovery from Coma and Disorders of Consciousness: A Preliminary Study. Am J Respir Crit Care Med. 2022;205(2):171–182. doi:10.1164/rccm.202105-1223OC

18. Maschke C, Duclos C, Owen AM, Jerbi K, Blain-Moraes S. Aperiodic brain activity and response to anesthesia vary in disorders of consciousness. NeuroImage. 2023;275:120154. doi:10.1016/j.neuroimage.2023.120154

19. Maschke C, Duclos C, Blain-Moraes S. Paradoxical markers of conscious levels: Effects of propofol on patients in disorders of consciousness. Frontiers in Human Neuroscience. 2022;16. Accessed October 6, 2022. https://www.frontiersin.org/articles/10.3389/fnhum.2022.992649

20. Colombo MA, Comanducci A, Casarotto S, et al. Beyond alpha power: EEG spatial and spectral gradients robustly stratify disorders of consciousness. Cerebral Cortex. Published online 2023:bhad031. doi:10.1093/cercor/bhad031

21. Lendner JD, Helfrich RF, Mander BA, et al. An electrophysiological marker of arousal level in humans. Elife. 2020;9:e55092. doi:10.7554/eLife.55092

22. Purdon PL, Pierce ET, Mukamel EA, et al. Electroencephalogram signatures of loss and recovery of consciousness from propofol. PNAS. 2013;110(12):E1142–E1151. doi:10.1073/pnas.1221180110

23. Alkhachroum A, Appavu B, Egawa S, et al. Electroencephalogram in the intensive care unit: a focused look at acute brain injury. Intensive Care Med. 2022;48(10):1443–1462. doi:10.1007/s00134-022-06854-3

24. Edlow BL, Fins JJ. Assessment of Covert Consciousness in the Intensive Care Unit: Clinical and Ethical Considerations. The Journal of Head Trauma Rehabilitation. 2018;33(6):424. doi:10.1097/HTR.0000000000000448

25. Duclos C, Norton L, Laforge G, et al. Protocol for the Prognostication of Consciousness Recovery Following a Brain Injury. Front Hum Neurosci. 2020;14. doi:10.3389/fnhum.2020.582125

26. Gramfort A, Luessi M, Larson E, et al. MEG and EEG data analysis with MNE-Python. Frontiers in Neuroscience. 2013;7. Accessed March 23, 2022. https://www.frontiersin.org/article/10.3389/fnins.2013.00267

27. Donoghue T, Haller M, Peterson EJ, et al. Parameterizing neural power spectra into periodic and aperiodic components. Nat Neurosci. 2020;23(12):1655–1665. doi:10.1038/s41593-020-00744-x

28. Pedregosa F, Varoquaux G, Gramfort A, et al. Scikit-learn: Machine Learning in Python. Journal of Machine Learning Research. 2011;12:2825–2830.

29. Jennett B, Bond M. Assessment of outcome after severe brain damage. Lancet. 1975;1(7905):480–484. doi:10.1016/s0140-6736(75)92830-5

30. Wilson J t. L, Pettigrew LE l., Teasdale GM. Structured Interviews for the Glasgow Outcome Scale and the Extended Glasgow Outcome Scale: Guidelines for Their Use. Journal of Neurotrauma. 1998;15(8):573–585. doi:10.1089/neu.1998.15.573

31. Rappaport M, Hall KM, Hopkins K, Belleza T, Cope DN. Disability rating scale for severe head trauma: coma to community. Arch Phys Med Rehabil. 1982;63(3):118–123.

32. Esnault P, Montcriol A, D’Aranda E, et al. Early neurological wake-up test in intubated brain-injured patients: A long-term, single-centre experience. Australian Critical Care. 2017;30(5):273–278. doi:10.1016/j.aucc.2016.10.002

33. Knaus WA, Draper EA, Wagner DP, Zimmerman JE. APACHE II: A severity of disease classification system. Critical Care Medicine. 1985;13(10):818. Accessed April 2, 2024. https://journals.lww.com/ccmjournal/abstract/1985/10000/APACHE_IIA_severity_of_diseas e_classification.9.aspx%E0%B8%A3%EF%A3%82

34. Vallat R. Pingouin: statistics in Python. Journal of Open Source Software. 2018;3(31):1026. doi:10.21105/joss.01026

35. Maschke C, Belloli L, Manasova D, Sitt JD, Blain-Moraes S. The role of etiology in the identification of clinical markers of consciousness: comparing EEG alpha power, complexity, and spectral exponent. Published online March 22, 2024:2024.03.20.24304639. doi:10.1101/2024.03.20.24304639

36. Kodaka M, Suzuki T, Maeyama A, Koyama K, Miyao H. Gender differences between predicted and measured propofol CP50 for loss of consciousness. Journal of Clinical Anesthesia. 2006;18(7):486–489. doi:10.1016/j.jclinane.2006.08.004

37. Leroy S, Major S, Bublitz V, Dreier JP, Koch S. Unveiling age-independent spectral markers of propofol-induced loss of consciousness by decomposing the electroencephalographic spectrum into its periodic and aperiodic components. Frontiers in Aging Neuroscience. 2023;14. Accessed September 27, 2023. https://www.frontiersin.org/articles/10.3389/fnagi.2022.1076393

38. Vuyk J, Oostwouder CJ, Vletter AA, Burm AGL, Bovill JG. Gender differences in the pharmacokinetics of propofol in elderly patients during and after continuous infusion. BJA: British Journal of Anaesthesia. 2001;86(2):183–188. doi:10.1093/bja/86.2.183

