## Supplementary Material for "EEG response during sedation interruption complements behavioral assessment following severe brain injury"

Content:

- Supplementary Methods
- Supplementary Table 1
- Supplementary Figure S1- S5

### Supplementary Methods:

***Reconfiguration score for induction of sedation***

All analysis presented in the main manuscript was performed on EEG recording during interruption of sedation (i.e., Phase 1-3). Phase 4 and 5 were available for 21 out of 42 patients. For those patients, the analysis was repeated using induction of sedation (i.e., Phase 3-5).

The change in each feature induced by the induction of propofol was defined as the difference of each features' average of the Phase 3 recording (sedation off) to the corresponding average of Phase 5 recording (sedation on). To evaluate whether this difference was caused by the induction of sedation rather than spontaneous fluctuations, we further assessed the proportion of datapoints post-induction (i.e., Phase 4 and 5) that reached values above or below one standard deviation of the sedation off (i.e., Phase 3) recording. The resulting value can be interpreted as a 'certainty score' with values close to 1 indicating high certainty that the respective feature changed after induction of sedation. Values of 0 indicate that observed changes in the mean are driven solely by natural fluctuations in the EEG over time.

The reconfiguration score of each feature was defined as the amount of change between sedation on and off, weighted by the above-described certainty score (i.e., amount of change * certainty of change). The reconfiguration score was calculated individually on mean, standard deviation and posterior-anterior ratio of each feature (i.e., total power, delta, theta, alpha, low beta, high beta, exponent and offset), yielding a total of 24 features (i.e., 8 mean, 8 standard deviation, 8 posterior-anterior ratio).

**Table S1:** Summary of included data

| | Phase 1 (on sedation) | Phase 2 (transition) | Phase 3 (off sedation) | Phase 4 (transition) | Phase 5 (on sedation) |
|---|---|---|---|---|---|
| Nr of recordings | 41 | 31 | 41 | 19 | 21 |
| Nr of epochs | 58.67 ± 14.69 | 58.41 ± 24.04 | 40.02 ± 17.99 | 44.45 ± 19.89 | 35.23 ± 12.25 |
| Nr of channels | 95.88 ± 8.79 | 95.53 ± 8.85 | 94.95 ± 6.8 | 94.6 ± 6.35 | 93.27 ± 6.27 |

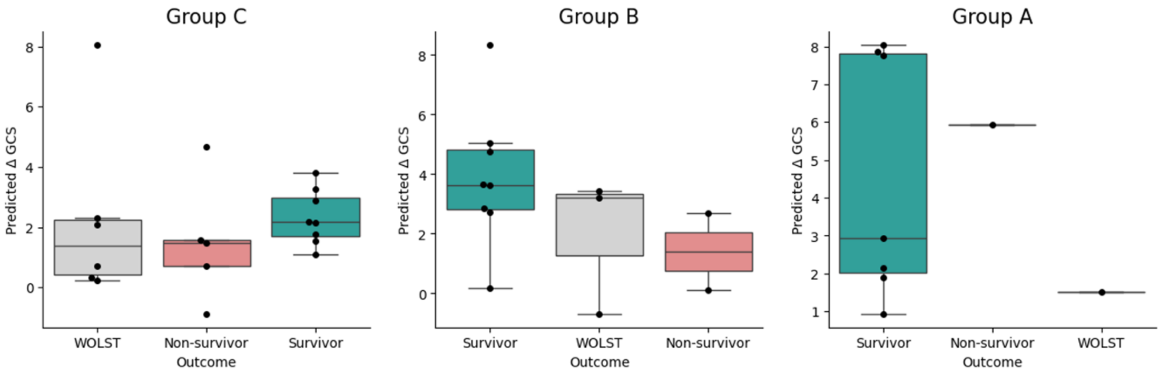


**Figure S1:** Model-predicted increase in Glasgow coma score (GCS) during neurological wake-up test. WOLST: withdrawn of life-supporting treatment. Each dot indicates a single patient. While patients in Group A showed a good behavioral response following interruption of sedation, the behavior of Group B and C was ambiguous or absent, respectively. WOLST: Withdrawal of life sustaining treatment.

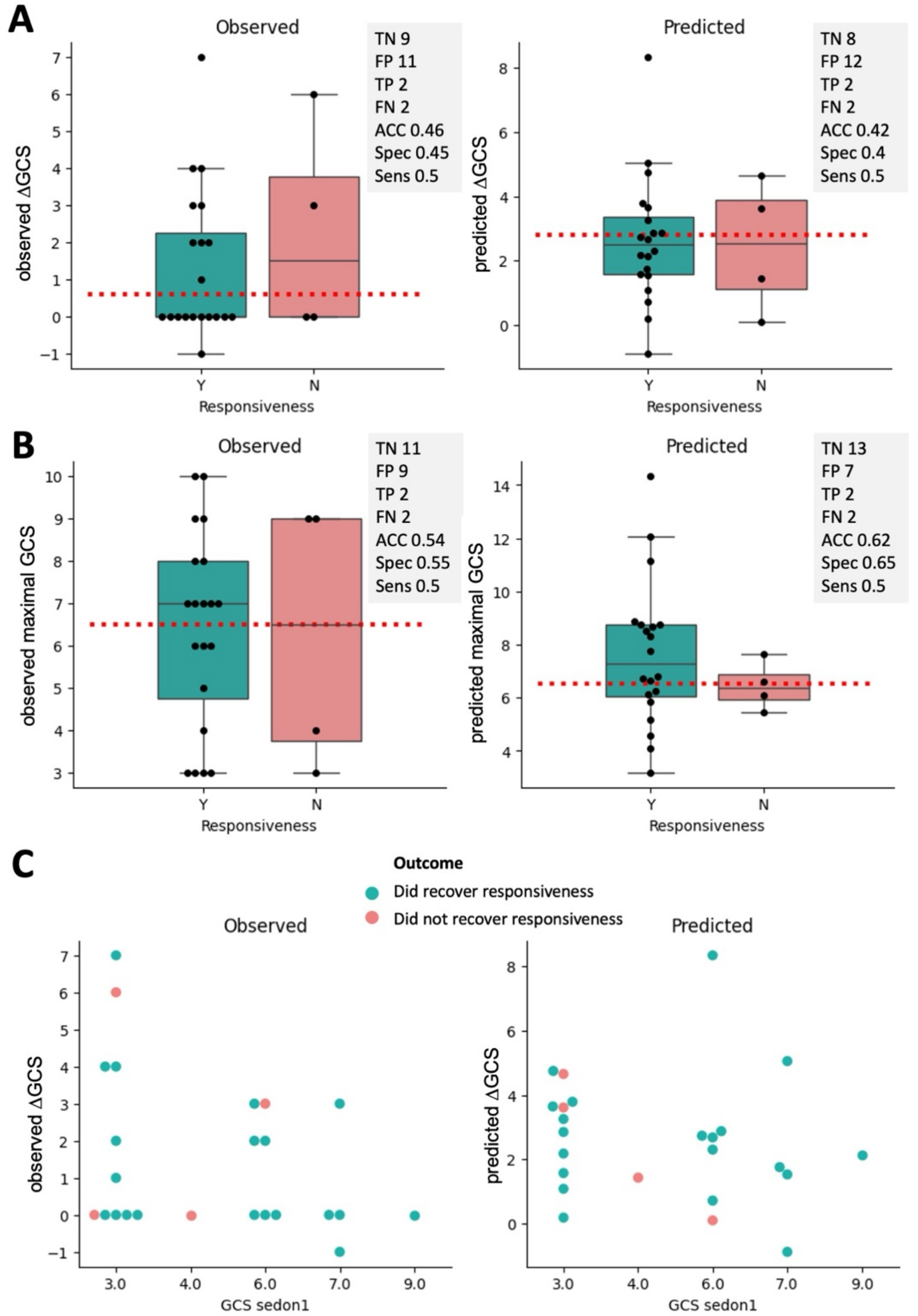


**Figure S2:** Comparison of observed and EEG-predicted behavioral response. **A)** Observed (left) and model-predicted (right) difference in Glasgow Coma Scale (GCS) during neurological wake- up test, split by patient's recovery of responsiveness (Y: Yes, N: No). **B)** Observed (left) and model-predicted (right) maximal Glasgow Coma Scale (GCS) during neurological wake- up test; The dotted line represents the value which best separates the favorable and unfavorable group. The grey box indicates performance matrices of group- separation based on this threshold (TN: true negative, FN: false negative, TP: true positive, FP: false positive, with positive indicating unfavorable outcome; ACC: accuracy, Spec: specificity, Sens: sensitivity). **C)** Visualization of group-separability based on the behaviorally observed response on sedation (i.e. GCS Phase 1) and the observed (left) and model-predicted (right) gain in GCS during neurological wake- up test.

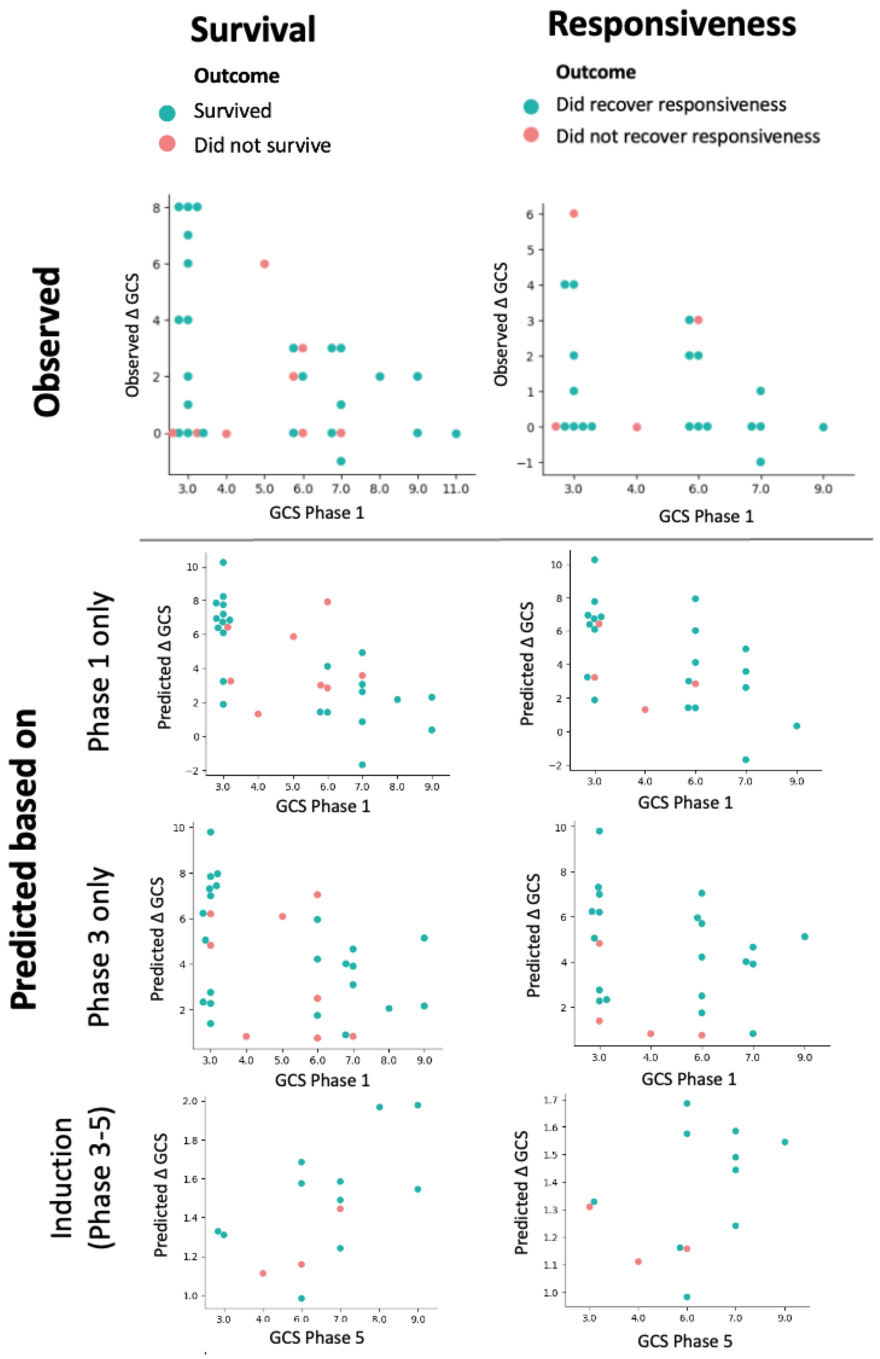


**Figure S3:** Comparison of observed (Top) and EEG-predicted (Bottom) behavioral response using features from different conditions: Phase 1 only) using feature mean over Phase 1 (sedation on), Phase 3 only) using feature mean over Phase 3 (sedation off), Induction) using feature as described in the supplementary methods extracted from Phase 3-5. Each dot represents one patient, colored by the respective outcome (green being favorable, red unfavorable).

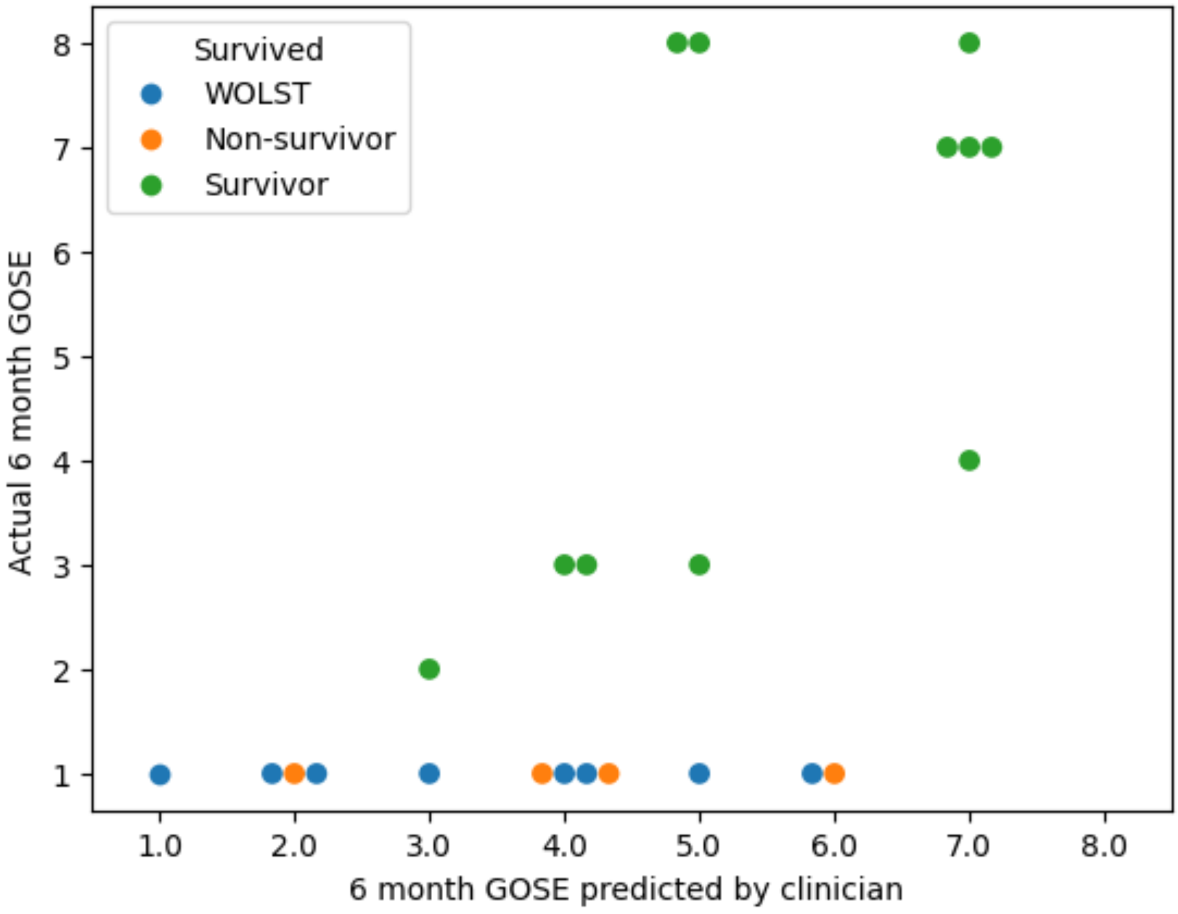


**Figure S4:** Actual versus clinician-predicted 6-month score on the Glasgow outcome scale extended (GOSE). Patients who were withdrawn from life sustaining treatments (WOLST) were not included in the analysis and are just present for purpose of visualization.

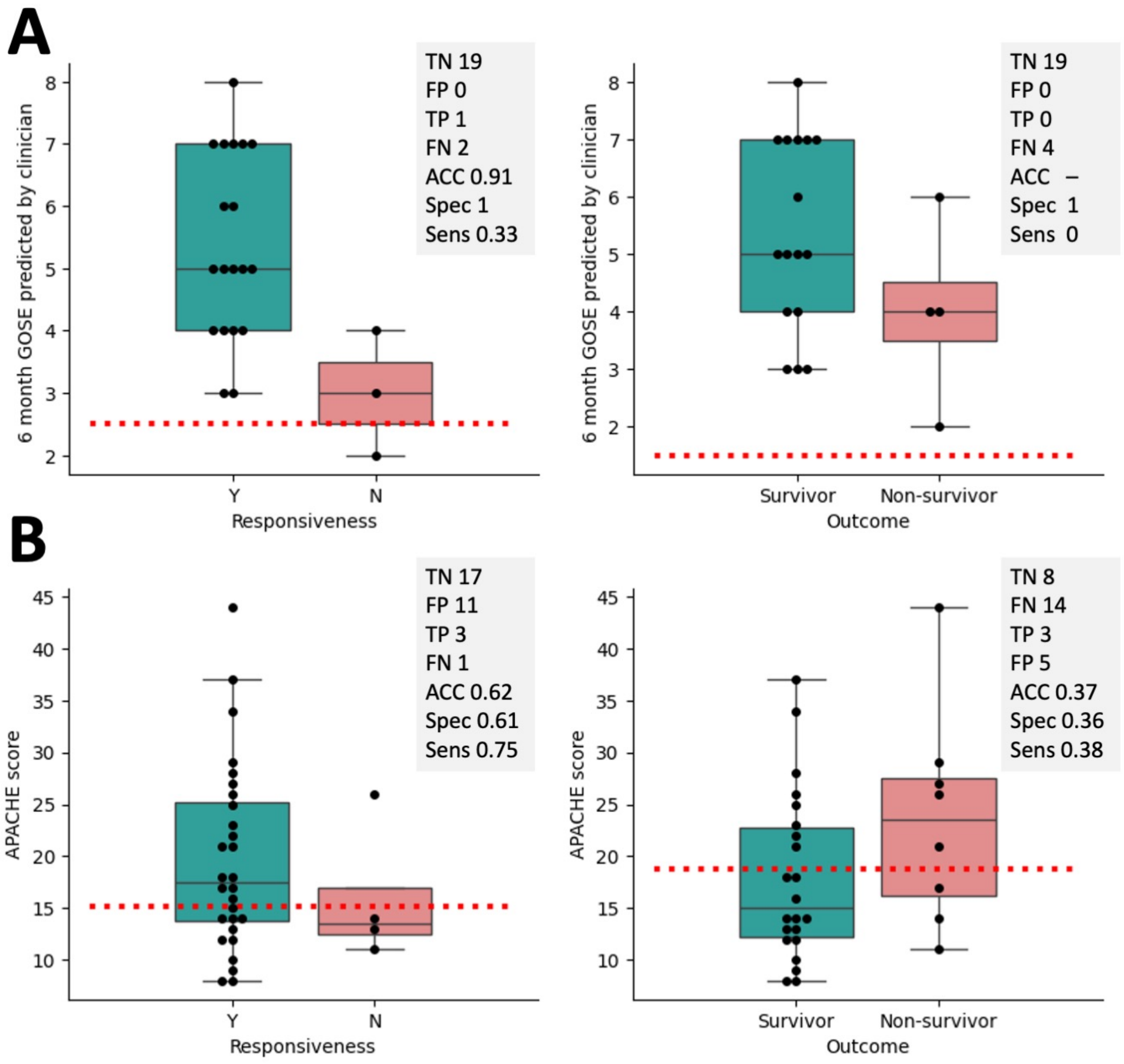


**Figure S5:** Prognostic value in clinician prediction and APACHE score. **A)** Prognostic performance of GOS-E score, predicted by clinicians, dotted lines represent a GOS-E above 2 (responsiveness) and GOSE above 1 to (survival). **B)** Prognostic performance of APACHE scores to prognosticate patient's recovery of responsiveness (Y: Yes, N: No) and survival. The dotted line represents the value which best separates the favorable and unfavorable group. The grey box indicates performance matrices of group-separation based on this threshold (TN: true negative, FN: false negative, TP: true positive, FP: false positive, with positive indicating unfavorable outcome; ACC: accuracy, Spec: specificity, Sens: sensitivity). Patients who were withdrawn from life sustaining treatments were not included in this analysis.
